# A new AI-assisted data standard accelerates interoperability in biomedical research

**DOI:** 10.1101/2024.10.17.24315618

**Authors:** Rodney Alan Long, Shannon Ballard, Syed Shah, Owen Bianchi, Lietsel Jones, Mathew J. Koretsky, Nicole Kuznetsov, Elise Marsan, Bryant Jen, Philip Chiang, Abhradeep Mukherjee, Cornelis Blauwendraat, Hampton Leonard, Dan Vitale, Kristin Levine, Sara Bandres-Ciga, Paige Jarreau, Patrick Brannelly, Caroline Pantazis, Laurel Screven, Kate Andersh, Alifiya Kapasi, John F. Crary, David Gutman, Brittany N. Dugger, Sarah Biber, Timothy Hohman, Faraz Faghri, Michael Griswold, Lana Sargent, Kendall van Keuren-Jensen, Andrew B. Singleton, Yang Fann, Mike A. Nalls, Hirotaka Iwaki

## Abstract

In this paper, we leveraged Large Language Models(LLMs) to accelerate data wrangling and automate labor-intensive aspects of data discovery and harmonization. This work promotes interoperability standards and enhances data discovery, facilitating AI-readiness in biomedical science with the generation of Common Data Elements (CDEs) as key to harmonizing multiple datasets. Thirty-one studies, various ontologies, and medical coding systems served as source material to create CDEs from which available metadata and context was sent as an API request to 4th-generation OpenAI GPT models to populate each metadata field. A human-in-the-loop (HITL) approach was used to assess quality and accuracy of the generated CDEs. To regulate CDE generation, we employed ElasticSearch and HITL to avoid duplicate CDEs and instead, added them as potential aliases for existing CDEs. The generated CDEs are foundational to assess the interoperability potential of datasets by determining how many data set column headers can be correctly mapped to CDEs as well as quantifying compliance with permissible values and data types. Subject matter experts reviewed generated CDEs and determined that 94.0% of generated metadata fields did not require manual revisions. Data tables from the Alzheimer’s Disease Neuroimaging Initiative (ADNI) and the Global Parkinson’s Genetic Program (GP2) were used as test cases for interoperability assessments. Column headers from all test cases were successfully mapped to generated CDEs at a rate of 32.4% via elastic search.The interoperability score, a metric for dataset compatibility to CDEs and other connected datasets, based on relevant criteria such as data field completeness and compliance with common harmonization standards averaged 53.8 out of 100 for test cases. With this project, we aim to automate the most tedious aspects of data harmonization, enhancing efficiency and scalability in biomedical research while decreasing activation energy for federated research.

## Introduction

Artificial intelligence/Machine learning (AI/ML) is a rapidly evolving tool with expanding applications in data processing and generalized data harmonization. This could be especially valuable for biomedical research areas such as Alzheimer’s disease and related dementias (ADRD) where AI/ML facilitated data harmonization can be a crucial step for new insights, but is not itself the end goal. “AI-readiness” is a major goal for federal health standards, particularly those relating to a digital National Institutes of Health (NIH) movement ^1^. Central to AI-readiness is establishing new standards for easy data discovery and interoperability across large organizations ^2^. Leveraging applied AI tooling via Large Language Models (LLMs) has been shown to accelerate the data wrangling process by an order of magnitude through automation of tedious aspects of data harmonization. This is especially true in health care where most data harmonization and common data element definitions are accomplished via what is essentially manual labor. Establishing these new data engineering and harmonization standards will also facilitate efficiency in future LLM-derived insights. Findability, Accessibility, Interoperability, and Reusability (FAIR) principles are essential to collaborative data science. Without the integration of common data elements (CDEs), performing federated or meta-analyses across data silos is not possible in clinical research ^3^.

Here, we discuss our AI-assisted Data Inventory and Verification Environment for the Research platform (DIVER), which can serve as a foundation for enhanced data discovery and interoperability standards in biomedical research. As a case study, we focus on data elements within the ADRD realm alongside heterogeneous clinical data. We utilize a user-friendly form structure for cataloging and indexing data and leverage applied-AI to build generative CDEs (GenCDEs) for tens of thousands of samples. We began by cataloging 72 studies, including both ADRD specific and generalist repositories. The DIVER tool enabled us to build and audit over 43,000 GenCDEs across the 31 of these studies, effectively reducing the data wrangling required for cross-study interoperability. This human-in-the-loop (HITL) approach where humans are partnered with AI tools, coupled with a foundationally open science approach, will advance the rate at which data silos can be made interoperable.

Automating the process of assessing data interoperability is key to AI-readiness in the biomedical space, particularly in light of recent developments relating to data locality requirements and federated learning. With data discovery and data wrangling making up a large portion of data science efforts, we expect that our approach will decrease the activation energy for researchers and research networks allowing time for more innovation. It will not only provide a growing collection of reference biomedical CDEs, but also a framework to harmonize disparate datasets rapidly, thereby advancing the speed of collaborative science.

## Methods

### Ethics statement

We exclusively used publicly accessible data without including any patient and/or participant-related or confidential information in AI applications. According to local regulations this is considered non-human subjects research and no ethics approval was required for this study. All procedures were conducted in accordance with the Declaration of Helsinki.

### Input data

To date we have cataloged and indexed 72 datasets for FAIR metadata inventory and discovery, of which 31 were included in the generation of subsequent CDEs (Supplemental Materials). FAIR metadata was manually cataloged for each study via Google Forms or within the DIVER app ^4^, summarizing content for our internal data inventory at NIH.

The DIVER app can be found here: https://diver-st-809832168532.us-central1.run.app and the DIVER API endpoint can be reached here: https://diver-api-809832168532.us-central1.run.app CDEs were generated by ingesting 31 datasets with clinical data (not primarily molecular datasets) along with the variables for definition from various sources. These included heterogeneous sources of data from ICD9 and ICD10 codes or other billing codes (including SNOMED and OMOP ontologies) as well as sparse data dictionaries from epidemiological studies. We then abstracted column headers that needed disambiguation via OpenAI’s 4th generation models. Column headers were defined variable names within a python data frame.

After cataloging studies, a subset of 6 datasets sharing participant-level data were analyzed as a proof-of-concept for matching our novel harmonized CDEs to existing data. This allowed us to calculate interoperability across data silos, as discussed below.

Please refer to **Figure 1** for a summary of the input data and the human-in-the-loop CDE system in general.

**Figure 1:**
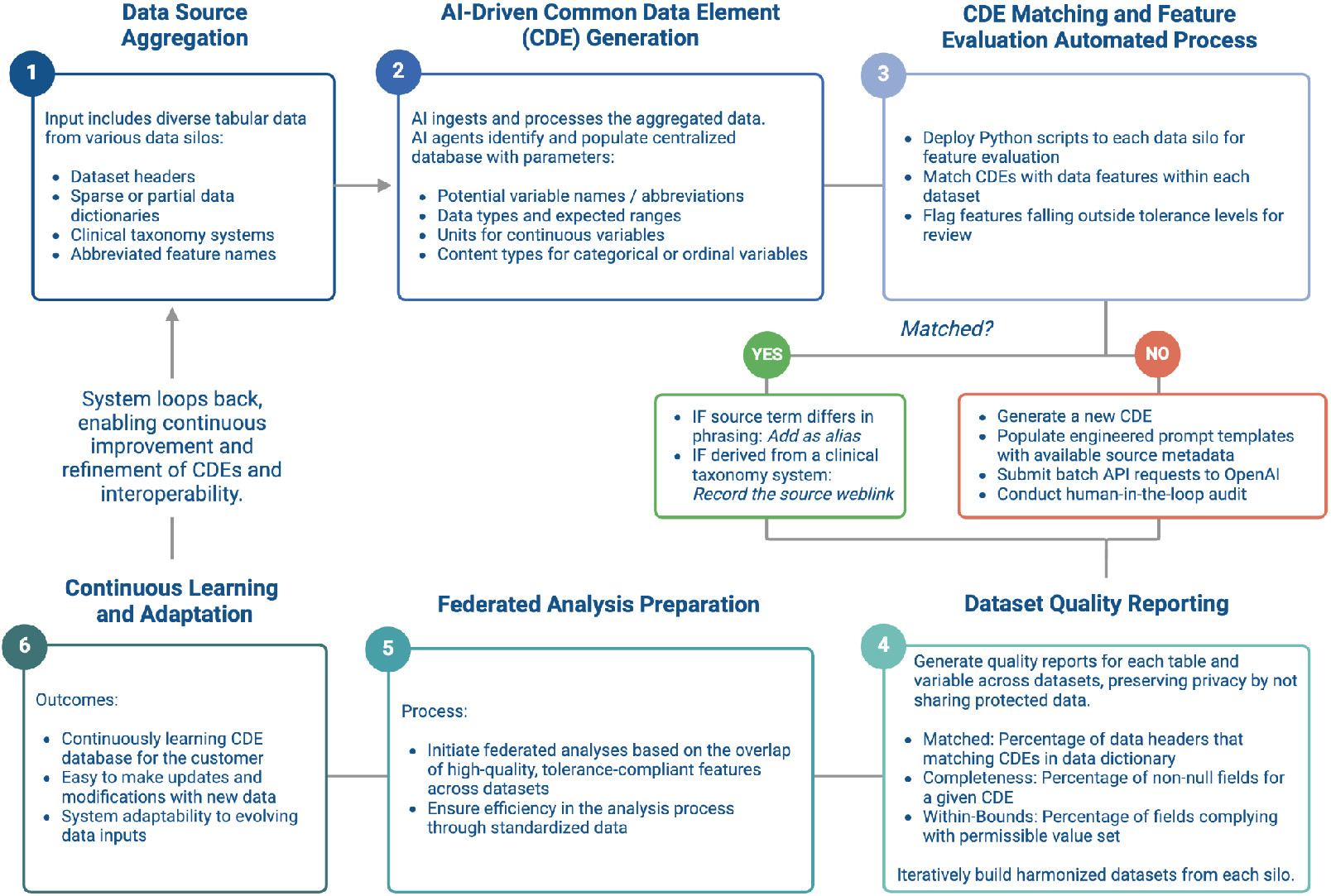
Workflow used to generate CDEs for interoperability assessments.

### AI-assisted CDE Generation

We employed an AI-assisted approach to generate CDEs from the ingested 31 datasets, leveraging iterative prompt engineering and template-based automation. Abbreviated data headers and other elements requiring definition were extracted from diverse data sources, see **Table 1**. These elements were processed through fourth generation OpenAI GPT models in a structured workflow to iteratively generate metadata elements, including alternative titles, abbreviations, permissible values, units of measure, preferred question text, and short descriptions. The complete analysis cost less than $1,000 USD.

**Table 1:** Sources used to generate CDEs. This table describes the 31 data sources used for CDE generation. ‘Origin’ refers to the input data source, including general ICD-10 coding from the NIH Clinical Center. ‘N elements’ represents the number of unique data objects that could potentially be converted into CDEs. ‘Hit rate’ indicates the percentage of metadata fields per CDE that did not require manual editing by the expert reviewer. ‘Source category’ specifies the data structure type used to build the CDEs. For web links to ingested content, please see the Supplemental Materials. A sparse dictionary may reflect a lack of extended titles (compared to abbreviated headers), standardization in data formatting and entry, and/or descriptions.

| Origin | n Elements | Hit Rate (%) | Source Category |
| --- | --- | --- | --- |
| BTRIS Clinical Observations | 12,437 | 100 | Established Coding System |
| CARD | 621 | 87 | Sparse Dictionary |
| Clinical Trials Database Form Legends | 1,578 | 87 | Sparse Dictionaries / Just Headers |
| Current Procedural Terminology - Healthcare Common Procedure Coding System (CPT – HCPCS) | 596 | 100 | Established Coding System |
| Gastrointestinal Symptom Rating Scale (GSRS) | 15 | 60 | Sparse Dictionary |
| Health Rhythms and Inferences | 99 | 91 | Sparse Dictionary |
| ICD | 16,989 | 100 | Established Coding System |
| Input Exercise Protocol | 6 | 100 | Sparse Dictionary |
| Intake Form Protocol | 282 | 33 | Sparse Dictionary |
| Kubio HRV | 27 | 80 | Sparse Dictionary |
| Metabolic Cart | 38 | 69 | Sparse Dictionary |
| MNPQ Protocol 20n0153 | 14 | 64 | Sparse Dictionary |
| Montreal Cognitive Assessment | 15 | 93 | Sparse Dictionary |
| National Institute of Mental Health Life Chart | 147 | 78 | Sparse Dictionary |
| National Library of Medicine (NLM) Endorsed | 137 | 100 | Established Coding System |
| National Library of Medicine (NLM) Qualified | 6,308 | 83 | Established Coding System |
| Neuropsychiatric | 1,105 | 80 | Sparse Dictionary |
| Ninehole Assessment | 7 | 94 | Sparse Dictionary |
| Non-Motor Symptom assessment scale for Parkinson's Disease (NMSS) | 100 | 73 | Sparse Dictionary |
| Nutrition Data System for Research | 120 | 100 | Sparse Dictionary |
| Populations Underrepresented in Mental illness Association Studies (PUMAS) | 1,143 | 60 | Just Headers |
| Pronutra | 12 | 100 | Sparse Dictionary |
| PsyToolKit | 10 | 100 | Sparse Dictionary |
| Resting State EEG | 101 | 100 | Sparse Dictionary |
| Rush Alzheimer's Disease Center | 79 | 90 | Sparse Dictionary |
| Safety Monitor Survey | 27 | 100 | Sparse Dictionary |
| Short Form Survey - 36 (SF-36) | 36 | 50 | Sparse Dictionary |
| Symp PsyTool Kit | 26 | 100 | Sparse Dictionary |
| Timed Up and Go (TUG) | 8 | 96 | Sparse Dictionary |
| Unified Parkinson's Disease Rating Scale (UPDRS) | 125 | 97 | Sparse Dictionary |
| MIMICEL (MIMIC-IV Event Log for Emergency Department) | 171 | 98 | Manual Inputs by Staff |
|  | **Total** | **Weighted Score** |  |
|  | 42,379 | 94 |  |

A range of parameters for the fourth generation GPT API models were used and are as follows: 100 Tokens, 0.5 - 0.7 Temperature. The token limit was more than generous. The 100 token limit was a consideration for a list of aliases that a CDE could be referred to and indeed some responses did reach that limit but on average only 13 tokens were required to complete a response. OpenAI states that a word is worth 1.25 tokens on average. Tokens can also be consumed by sub words and trailing spaces.

Custom prompt engineering was performed by inserting generated metadata fields using the Jinja templating engine^4^ for the automation and standardization of prompts sent to the AI models. For instance, if the CDE source was an abbreviated data header, such as ‘wbc_count’, it was inserted into the prompt template instructing the GPT model to expand it into a full, descriptive data element title. This expanded title was inserted into subsequent prompt templates for further metadata generation. The next step typically involved generating a short description of the data element, followed by creating question text to retrieve the data of interest. Once the data title and short description were available, they were inserted into a template to classify the data element by type (e.g., numerical continuous, categorical ordinal, or binary).

For numerical data types, the generated data title, type, and question text were inserted into a template that tasked the GPT model with assigning a unit of measure, where applicable. The unit of measure and question text were then used to generate a set of permissible values denoting acceptable content and ranges per CDE.

### Human-in-the-Loop

Throughout the process of generating CDEs, a human-in-the-loop played a crucial role in auditing the AI-generated outputs. Typically, a descriptive title was the first item generated for a CDE from an abbreviated header. The human expert reviewer assessed whether the source was interpreted correctly and determined whether detail and specificity of the generated title was suitable for generating additional metadata. The human-in-the-loop had the discretion to adjust the prompt order or include additional metadata elements based on the complexity and quality of the source material. As an example, the following header was encountered: *IQ_CS_SCD_HR_EX*. This is complex as you have five abbreviations in a single header. The quality could be considered poor due to the terms being severely abbreviated and commonly applied to other concepts. Initially, GPT interpreted it as *Interviewee’s Clinical Survey Weeknight Exercise Hours*. The correct expansion in this case was *Intake Questionnaire - Child Survey - School Day - Hours of Exercise*. In such situations, it is important to add additional context so that GPT may correctly interpret the abbreviations. For this example, it was important to include that the table contained data from an intake questionnaire for school children to assess and compare the hours of physical activity on school days vs weekends.

This hybrid approach—combining AI-driven metadata generation with expert oversight—helped ensure that the resulting CDEs were both accurate and contextually relevant. Once generated and reviewed, the CDEs were integrated into the centralized data dictionary hosted by DIVER, which serves as a large-scale repository for standardized elements. This AI-assisted process significantly accelerated the creation of CDEs, reducing the time required to establish data harmonization across studies.

We implemented a human-in-the-loop system to ensure the quality and accuracy of the AI-generated Common Data Elements (CDEs). Subject Matter Experts (SMEs) including two clinical data scientists (RAL, LJ), one medical doctor (HI) and two biomarker scientists (KvJ and EM) manually audited the CDEs, focusing on criteria such as relevance, accuracy, and adherence to existing standards. This auditing process was essential in verifying that the AI-generated outputs were suitable for biomedical research and met the rigorous standards required for data harmonization.

The uniformity and comprehensiveness of the generated CDEs facilitated a streamlined evaluation process, enabling rapid review. A single SME could audit over 4,000 CDEs per week at peak efficiency. This could vary widely with subject matter; an average efficiency is estimated at 1,500 CDEs per week. Internal tests were compared to current best practices including heavy manual efforts for CDE curation generating less than 400 CDEs in a week of full-time effort from one SME. The manual auditing step was critical in refining the CDEs by identifying errors or inconsistencies requiring intervention.

During the auditing process, the generated CDEs were scored based on the percentage of fields that required manual revision. A manual revision was deemed necessary when metadata was inconsistent with the style and information content of previously discussed ontologies and the 184 endorsed NLM CDEs int he public domain. Some examples of this included solicitation of string values by question text while the permissible expected values were numerical or when expanded abbreviations were inaccurate (see Supplement). Cases where minor modifications were needed—such as removing prefixes or labels automatically appended by the GPT model—were not considered full manual interventions as they could be corrected through bulk updates. For example, despite explicit instructions to omit labels, the GPT model often included phrases like “Permissible Values:” preceding the value set. These corrections were handled efficiently without extensive manual input and were not counted among the manual edits, but rather part of post-processing scripts.

### Learning System and ElasticSearch

To expand our Common Data Elements (CDEs) repository, we applied an approach that combined automated processes and advanced search technologies across various datasets. Python scripts were deployed across datasets (treated as silos) to evaluate features and match them with existing CDEs to avoid adding duplicates. This allows the system to learn which new CDEs may need development based on simple matching via BM25 ElasticSearch ^5^.

We integrated Elasticsearch to enhance the matching process. Elasticsearch’s advanced search capabilities allowed us to efficiently retrieve and suggest relevant CDEs from our growing repository, optimizing the matching of CDEs with data dictionaries and column headers from research initiatives not part of the CDE generation process. These included the Alzheimer’s Disease Neuroimaging Initiative (ADNI), the Global Parkinson’s Genetic Program (GP2), and pulled records from MIMIC-IV Event Log (MIMICEL), see **Table 1**.

Our Elasticsearch configuration utilized a two-shard, one-replica^6^ setup to balance performance and redundancy. Custom analyzers were developed to accurately tokenize varying case formats such as snake_case and CamelCase. To refine search relevance, we applied differential field weightings: aliases and title fields were weighted at ^4, short_description at ^3, preferred_question_text at ^2, and unit_of_measure at ^1. These weights dictate the influence of each field on the relevance score, with a weighting of ^4 contributing four times more than a weighting of ^1. The default BM25 parameters were employed (k1 = 1.2, b = 0.75) with fuzziness set to ‘auto’, which has an implicit threshold for matching.^7^ The fuzziness auto parameter determines matches as follows: an edit distance of 0 for strings of 1 to 2 characters, an edit distance of 1 for strings between 3 and 5 characters, and an edit distance of 2 for strings with more than 5 characters. It is important to note that the threshold is applied after the tokenization of terms.^8^ With this approach, a robust and flexible search was performed across our dataset.

If features matched existing CDEs but the source term differed in phrasing, it was recorded as an alias to maintain consistency within the CDE repository. Additionally, when CDEs were derived from clinical taxonomy systems, we documented the source weblink to maintain traceability and provide context.

When features did not align with existing CDEs, we initiated the creation of new CDEs. This process involved populating engineered prompt templates with available source metadata and submitting batch API requests to OpenAI. Pricing varies by model and is reduced by using batch API ^9^. The AI-generated metadata for these new CDEs was then reviewed through a human-in-the-loop process, which was crucial for verifying accuracy and relevance.

### Interoperability Scoring

An Interoperability Scoring system was implemented to evaluate the effectiveness of CDE utilization and compatibility across datasets based on three criteria: **Matching, Completeness**, and **Compliance**. This provides analysts an easily interpretable metric to make decisions regarding data inclusion and could be used as a filtering criterion. It also permits the prioritization of resources and estimation of manual labor needed for data harmonization efforts.

### Matching

was evaluated based on the ability to map column headers from external sources to CDEs within our centralized data dictionary using ElasticSearch. The descriptive and contextual metadata fields of generated CDEs were used for this task. The fields we included were the following: variable name, title, short description, preferred question text, and aliases. The percentage of external column headers correctly matched to the CDEs in the repository was calculated. If a data point could not be matched to an existing CDE, an expert reader evaluated whether a relevant CDE exists but was not matched or if an entirely new CDE should be generated.

### Completeness

was evaluated by determining the percentage of non-missing data fields for successfully mapped columns. Missing values can limit the ability to map data between columns, as these gaps mean some data points lack corresponding values for associative analysis.

### Compliance

was examined to ensure the values within non-null data points conformed to the permissible value sets associated with our generated CDEs. This criterion verified that data values within matched columns were recognized and fit within the acceptable ranges or categories as defined for each CDE. For continuous variables, the upper and lower ranges of values from the CDEs were used. For categorical variables (including ordinal variables redefined as categorical), compliance was based on the ability for a second round of BM25 ElasticSearch on parameters mirroring the initial matched phase.

These criteria collectively provided a comprehensive evaluation of dataset interoperability with the CDE repository, reflecting both the accuracy of data matching and the quality of data completeness and adherence to reporting conventions. The **Interoperability Score** is the percentage of values within a data field matched to a CDE that were not missing and within compliance which is then averaged per data frame.

The resulting Interoperability score formula is: *([% matching columns] + [% completeness of matched columns] + [% compliance of complete entries in matched columns])/3*. This will likely change as CDE databases expand and matching tools improve with time. Please see **Figure 2** for a summary of interoperability.

**Figure 2:**
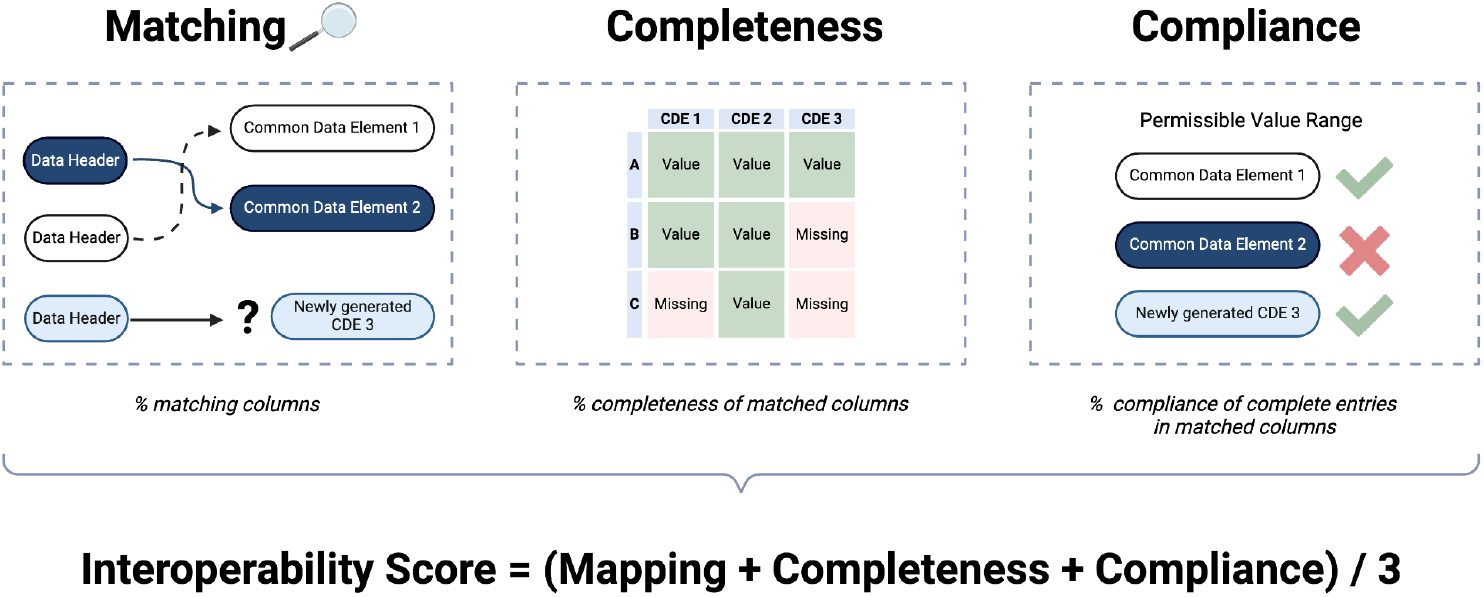
Summary of interoperability scoring.

## Results

### AI-accelerated generation of experimental and clinical CDEs

**Table 1** summarizes the effectiveness of using applied-AI to accelerate the creation of CDEs. This process automates much of the tedious foundational work of looking up variable names, aliases and permissible values as well as formatting data outputs. This efficiency saves the experts in the human-in-the-loop system immense amounts of time compared to de-novo construction.

The AI-driven approach significantly accelerated the generation of CDEs from data headers, medical coding systems, and spare or established data dictionaries. Across all data sources, GPT-4 and GPT-4o achieved a weighted accuracy rate of 94.0% in generating CDEs, with accuracy defined as the percentage of generated fields that required no manual revisions and weighted by averaging across CDE metadata fields rather than across data sources.

84.6% (35,853) of CDEs were derived from established clinical classification systems, while 15.4% (6,508) were generated from unstructured sources such as data headers and sparse data dictionaries. The overall unweighted accuracy, which averaged accuracy by source rather than per element, was 85.9%. When excluding established clinical classification systems, the unweighted hit rate for CDEs generated from unstructured sources was 83.8%.

These results demonstrate the efficacy of AI models in automating CDE generation, particularly when dealing with structured clinical systems. Variability in hit rates across data sources suggests that further optimization may be needed for unstructured or sparse data inputs.

### ElasticSearch matching of data fields to CDEs

ElasticSearch was employed to enhance the efficiency of matching existing data fields with appropriate CDEs. By leveraging ElasticSearch’s powerful text-based search capabilities, we were able to compare field names, descriptions, and associated metadata with a pre-existing repository of CDEs.

Headers from sources external to the material used to construct the current set of CDEs successfully matched in 25.8% of cases. These headers were pulled from the Alzheimer’s Disease Neuroimaging Initiative (ADNI) and the Global Parkinson’s Genetics Program (GP2), our two test cohorts ^11,12^. We also tested against clinical inputs including manually entered free-text notes from MIMIC-IV Event Log (MIMICEL) from the NIH Clinical Center ^13^.

At the time of writing, the data dictionary contained a subset of clinically relevant ICD codes. A total of 13,425 unique ICD codes from MIMICEL dataset were correctly matched at a rate of 48.9%. From the data set, 27,660 unique terms in clinician notes were extracted from recorded chief complaints, of which 4.4% were able to be matched to existing CDEs within the data dictionary. This is summarized in **Table 2**.

**Table 2:** Mapping test results. Description of the six tables used to test matching capabilities. Data table refers to a specific table from a set. Term source indicates whether terms were extracted from data headers or a specific column. ‘N terms’ indicates how many unique terms were extracted from the table. This includes data headers, ICD codes, and the documented chief complaints of a patient. The ‘confirmed match’ reports the percentage of returned matches that were verified.

| Table | Term Source | *n* Terms | Confirmed Match (%) |
| --- | --- | --- | --- |
| ADNI Merge | column header | 116 | 12.1 |
| ADNI MOCA | column header | 56 | 17.8 |
| ADNI ADSX | column header | 41 | 34.1 |
| GP2 | column header | 421 | 29.4 |
| MIMICEL Diagnosis | icd column | 13,425 | 48.9 |
| MIMICEL Triage | chief complaint column | 27,660 | 4.4 |

### Assessing interoperability for data silos

The interoperability of a dataset was assessed based on three criteria. The first being *Matching*; the percentage of data headers accurately matched to existing CDEs within the data dictionary. The second criteria was *Completeness*; the percentage of non-null fields for an accurately matched data header. The third criteria examined was *Compliance*; the percentage of non-null fields containing values that were recognized and fit within the permissible value set. As previously described, these three measures were then combined into a comprehensive *Interoperability score*. We summarized exploratory analyses of interoperability in our test datasets detailed in **Table 3**.

**Table 3:** Interoperability test results. Interoperability results based on three tables. ‘N columns’ indicate the total number of columns for the table. Matching reports the percentage of columns that were accurately matched to a CDE within our repository. Completeness reports the percentage of non-null fields for a matched column. Compliance is the percentage of non-null values that were within the bounds of our defined permissible value set. Interoperability score is the average score of the three criteria.

| Table | *n* Columns | Matching (%) | Completeness (%) | Compliance (%) | Interoperability Score |
| --- | --- | --- | --- | --- | --- |
| GP2 | 421 | 29.4 | 16.4 | 94.6 | 46.8 |
| ADNI Merge | 116 | 12.1 | 53.8 | 100 | 55.3 |
| ADNI MoCA | 56 | 17.8 | 90.8 | 75 | 61.2 |
| ADNI ADSXLIST | 41 | 34.1 | 100 | 21.4 | 51.8 |

While these data sets have established dictionaries, they were not ingested into our CDE repository prior to testing in order to assess real world applications as test data.

A sparse GP2 dataset from multiple clinical sites was used for testing. Of 421 data headers, 29.4% were confirmed as correct matches, 16.4% of the matched data fields contained non-missing values, and 94.6% of those values were recognized and in compliance with the permissible value set. This suggests that data that can be matched and is complete is generally within tolerances and nearly analysis ready. Although sparsity led to a total interoperability score of 46.8%.

ADNI Merge is a table that represents the consolidation of multiple ADNI research phases. Of the 116 data headers 12.1% were correctly matched to CDEs in our repository. Of those matches, 53.8% of the fields were populated and 100% of the values adhered to the set permissible values. A interoperability score of 52.7% was achieved. ADNI MoCA contains data related to the Montreal Cognitive Assessment. Of the 56 column headers, 17.8% were matched to existing CDEs within the repository. Of the matched data headers, 90.8% of the fields were populated and 75% of them complied with set permissible values. The overall interoperability score was calculated to be 61.2%. ADNI ADSXLIST refers to the Alzheimer’s Diagnosis and Symptom Checklist. Of the 41 data headers, 34.1% matched established CDEs within our repository. Matched columns were 100% populated and 21.4% of the values complied with set permissible values. The calculated interoperability score for this table was 52.7%.

These estimates across ADNI sub-studies illustrate how different aspects of the data harmonization process and the inherent difficulty of harmonization even within studies can be quantified by the interoperability scoring system described here. Low matching rates to existing CDEs is the primary problem for automated harmonization and off-the-shelf interoperability. As this system grows, this should improve.

### Time savings for novel CDE generation

This proof of concept for AI-assisted human-in-the-loop data harmonization illustrates the time-saving utility of this technology. AI used as a tool for CDE generation, matching, value look-ups and interoperability saves time on an order of magnitude compared to traditional manual efforts (less than 400 versus over 4,000 CDEs per week at peak efficiency per SME). Automating the tedious aspects of data harmonization and the logistical benefits of interoperability score under SME oversight will serve as the key to AI readiness in health research.

## Discussion

This work underscores the transformative potential of AI-assisted methodologies in advancing biomedical research, particularly in the realm of data interoperability and the generation of CDEs. By employing the most current AI models, specifically OpenAI’s GPT-4 and GPT-4o, in conjunction with human oversight, we have significantly accelerated the process of CDE generation and data standardization across a wide array of data silos. We have designed a system to automate the most tedious aspects of data harmonization.

### Interpretation of Results

The AI-driven approach achieved a weighted accuracy rate of 94.0% in generating CDEs, significantly reducing the amount of manual work required and enabling the rapid generation of large quantities of CDEs—far faster than could be achieved through manual methods. This high accuracy, with 84.6% of CDEs derived from established clinical classification systems, demonstrates the method’s robust performance when applied to structured data sources. Structured data, characterized by well-defined variables and consistent formats, allows AI models to more effectively generate and standardize CDEs that are uniformly formatted, leading to quicker and more reliable outcomes.

In contrast, the hit rates were slightly lower when applied to unstructured data sources, such as sparse data dictionaries or abstracted data headers (83.8% unweighted and 90.8% weighted). Unstructured data, which often needs more consistent formatting and can be more ambiguous, presents greater challenges for AI models. These challenges highlight the need for further optimization, potentially through more advanced machine learning techniques or enhanced context awareness, to improve the model’s ability to interpret and standardize unstructured inputs accurately.

Integrating ElasticSearch into the CDE matching process underscores the critical role of advanced search technologies in biomedical data management. While the success rate in matching headers from external sources was moderate (25.8%), the ability to correctly map 48.9% of ICD codes from a large-scale dataset like MIMIC-IV demonstrates the robustness of this approach for clinical data. However, the lower matching rates observed for unstructured terms (4.4%) from recorded chief complaints suggests that enhancing the natural language processing capabilities of the system or providing additional contextual information could improve the accuracy and consistency of CDE generation from unstructured or semi-structured data sources.

### Limitations and Future Directions

Despite these promising results, several limitations were identified. One of the primary challenges encountered was the model’s performance with sparse or novel data sources. The lack of contextual information in the data demonstrates an increased need for manual review in some cases. The absence of detailed metadata, such as question text or permissible values, increased the difficulty for the AI to interpret heavily abbreviated elements accurately, leading to occasional misinterpretations. We also note that there is a major English language bias in this analysis and further testing will be needed to account for the introduction of AI powered translations.

With the rapid advancement of GPT models, we have also seen the expansion of context windows and reduced token cost. Our approach utilized a zero-shot prompting which means we did not provide specific examples, nor did we provide feedback or corrections. In the future, it could be possible to provide an entire subset of CDEs related to submitted source material as examples. This could potentially increase quality and consistency. Perhaps a future GPT model would be able to recognize potential redundancies and suggest CDEs to fill gaps in coverage.

Institutional review board

Fine-tuning the AI model on domain-specific datasets could be a valuable next step to address these challenges. Fine-tuning allows the model to more precisely adjust to the specific context of the data it is processing, improving its ability to generate accurate CDEs from unstructured, semi-structured, or novel data sources. This process could reduce the need for manual oversight and increase the system’s efficiency, particularly in specialized fields where data may not conform to widely established standards.

The project also underscored the importance of effective data preprocessing. Enhancements in preprocessing techniques could mitigate some of the challenges associated with unstructured data. For instance, developing more sophisticated methods to contextualize ambiguous terms and applying organizing principles tailored to the specific characteristics of the data could further reduce the potential for misinterpretation by the model. These improvements would maintain the system’s high accuracy across a broader range of data types.

Another area for future improvement involves the human-in-the-loop process. While essential for ensuring the quality of the generated CDEs, the reliance on manual review introduces a bottleneck in scalability Automating certain aspects of the quality assurance process, such as implementing machine learning-based checks to flag potential errors for human review, could streamline this step. This would enhance the system’s scalability, making it more suitable for larger or more complex datasets without compromising accuracy.

## Conclusion

This project has demonstrated the significant potential of AI-driven solutions in the field of biomedical data standardization. The high accuracy rates achieved, particularly with structured data, highlight the system’s capability to reduce the manual labor involved in CDE generation, offering substantial improvements in both speed and efficiency. While there are areas for further refinement—particularly in handling unstructured or novel data—the project’s overall success provides a strong foundation for automated CDE generation. By incorporating fine-tuning, enhancing preprocessing techniques, and improving scalability through automation, this system could become an even more powerful tool in standardizing biomedical data, ultimately advancing research efforts across diverse scientific domains.

## Data Availability

Open science resources
Common data elements (CDEs) are housed for public access as part of a large data harmonization initiative at NIH. They are accessible through the DIVER (Data Inventory and Verification Environment for Research) web app that can be found here. The API endpoint can be found here.
The CDEs are accessible to the public without login. Just click the 'General Users' identity tab, navigate to 'query data' on the CDE tab. There are a total of over 43,000 human-in-the-loop validated CDEs available for data harmonization efforts on the DIVER web app (potentially the largest public repository of this sort), some of these relate to clinical data, EMR and REDCap related ontologies. To limit to curated collections for sets of related CDEs, please filter by 'collections' then select collections of interest in the drop down.
Example of DIVER API Call in Supplemental.
Table linking to data sources also in Supplemental.

## Author Affiliations

See above.

## References

1. Fagnan, K. et al. Data and Models: A Framework for Advancing AI in Science. https://www.osti.gov/biblio/1579323 (2019) doi:10.2172/1579323.

2. Scheffler, M. et al. FAIR data enabling new horizons for materials research. Nature 604, 635–642 (2022).

3. Sheehan, J. et al. Improving the value of clinical research through the use of Common Data Elements. Clin. Trials (2016) doi:10.1177/1740774516653238.

4. Jinja Documentation. Jinja https://jinja.palletsprojects.com/en/3.1.x/.

5. Rivas, A. R., Iglesias, E. L. & Borrajo, L. Study of query expansion techniques and their application in the biomedical information retrieval. ScientificWorldJournal 2014, 132158 (2014).

6. Reading and Writing documents - ElasticSearch https://www.elastic.co/guide/en/elasticsearch/reference/current/docs-replication.html.

7. Relevance Tuning Guide, Weights and Boosts. https://www.elastic.co/guide/en/app-search/current/relevance-tuning-guide.html.

8. Fuzziness. https://www.elastic.co/guide/en/elasticsearch/guide/current/fuzziness.html.

9. Website. https://openai.com/api/pricing/.

10. World Health Organization. The International Statistical Classification of Diseases and Health Related Problems ICD-10: Tenth Revision. Volume 2: Instruction Manual. (World Health Organization, 2004).

11. Petersen, R. C. et al. Alzheimer’s Disease Neuroimaging Initiative (ADNI): Clinical characterization. Neurology 74, 201 (2010).

12. Global Parkinson’s Genetics Program. GP2: The Global Parkinson’s Genetics Program. Mov. Disord. 36, 842–851 (2021).

13. Goldberger, A. L. et al. PhysioBank, PhysioToolkit, and PhysioNet: components of a new research resource for complex physiologic signals. Circulation 101, E215–20 (2000).

14. OpenAI Platform. https://platform.openai.com/tokenizer.

