## Supplemental Material for "A new AI-assisted data standard accelerates interoperability in biomedical research" (from another version of this preprint: version 1, 2024-10-17)

### Open science resources

Common data elements (CDEs) are housed for public access as part of a large data harmonization initiative at NIH. They are accessible through the DIVER (Data Inventory and Verification Environment for Research) web app that can be found here. The API endpoint can be found here.

The CDEs are accessible to the public without login. Just click the “General Users” identity tab, navigate to “query data” on the CDE tab. There are a total of over 43,000 human-in-the-loop validated CDEs available for data harmonization efforts on the DIVER web app (potentially the largest public repository of this sort), some of these relate to clinical data, EMR and REDCap related ontologies. To limit to curated collections for sets of related CDEs, please filter by “collections” then select  collections of interest in the drop down.

Example python API calls are below:

```
from requests.exceptions import HTTPError
##### INLINE README
# Return value is a list of list:
# res: final result can be retrieved from hits = res['hits']['hits']
# WHERE:
#  hits is a list and final response can be assembled by iterating over this list
# response -> hit['_source'] for each hit in hits
###############
new_qa=pd.read_csv("es_qa.csv")
queries = list(new_qa['question'])
link="https://diver-api-809832168532.us-central1.run.app/run_query"
top_k="100"
for idx, query in enumerate(queries):
    if idx <2: #restric to 2 queries for demo purpose
        d = {"query": f"{query}", "top_k": f"{top_k}"}

        print(f'd: {d}')
        print(f'json.dumps(d): {json.dumps(d)}')
        try:

            r = requests.post(f"{link}", data=json.dumps(d))

            r.raise_for_status()
            res=r.json()
```

```
        final_ans = []
        for hit in res['hits']['hits']:

            if hit['_score']>8:
                r=hit['_source']
                final_ans.append(r)
        res_df=pd.DataFrame(final_ans)
        if res_df.shape[0]>0:
            print(f"query: {query} \nres_df: {res_df}")
    except HTTPError as http_err:
        print(f'HTTP error occurred: {http_err}')
    except Exception as err:
        print(f'Other error occurred: {err}')
```

### Weblinks to Data Sources

| **Origin** | **Weblink** |
|---|---|
| BTRIS Clinical Observations | https://btris.nih.gov/ |
| CARD | https://card.nih.gov/data-resources/access-data |
| Clinical Trials Database Form Legends | https://ctdb.nichd.nih.gov/ |
| Current Procedural Terminology - Healthcare Common Procedure Coding System (CPT – HCPCS) | https://www.cms.gov/medicare/regulations-guidance/physician-self-referral/list-cpt-hcpcs-codes |
| Gastrointestinal Symptom Rating Scale (GSRS) | https://pubmed.ncbi.nlm.nih.gov/3123181/ |
| Health Rhythms and Inferences | https://www.healthrhythms.com/ |
| ICD | https://www.cms.gov/medicare/coding-billing/icd-10-codes |
| Input Exercise Protocol | |
| Intake Form Protocol | |
| Kubio HRV | https://www.kubios.com/ |
| Metabolic Cart | |
| MNPQ Protocol 20n0153 | |
| Montreal Cognitive Assessment | https://mocacognition.com/ |
| National Institute of Mental Health Life Chart | https://pubmed.ncbi.nlm.nih.gov/9368200/ |

| | |
|---|---|
| National Library of Medicine (NLM) Endorsed | https://cde.nlm.nih.gov/home |
| National Library of Medicine (NLM) Qualified | https://cde.nlm.nih.gov/home |
| Neuropsychiatric | |
| Ninehole Assessment | https://www.ncbi.nlm.nih.gov/pmc/articles/PMC5405844/ |
| Non-Motor Symptom assessment scale for Parkinson's Disease (NMSS) | https://www.movementdisorders.org/MDS/MDS-Rating-Scales/Non-Motor-Symptoms-Scale-for-Parkinsons-Disease-NMSS.htm |
| Nutrition Data System for Research | https://www.ncc.umn.edu/ndsr-database-page/ |
| Populations Underrepresented in Mental illness Association Studies (PUMAS) | https://loeslab.dgsom.ucla.edu/projects/pumas |
| Pronutra | https://www.viocare.com/pronutra.html |
| PsyToolKit | https://www.psytoolkit.org/ |
| Resting State EEG | |
| Rush Alzheimer's Disease Center | https://www.radc.rush.edu/docs/var/standardDatasetVariables.htm |
| Safety Monitor Survey | |
| Short Form Survey - 36 (SF-36) | https://www.rand.org/health-care/surveys_tools/mos/36-item-short-form.html |
| Symp PsyTool Kit | https://www.psytoolkit.org/ |
| Timed Up and Go (TUG) | https://www.cdc.gov/steadi/media/pdfs/STEADI-Assessment-TUG-508.pdf |
| Unified Parkinson's Disease Rating Scale (UPDRS) | https://www.movementdisorders.org/MDS/MDS-Rating-Scales/MDS-Unified-Parkinsons-Disease-Rating-Scale-MDS-UPDRS.htm |
| MIMICEL (MIMIC-IV Event Log for Emergency Department) | https://physionet.org/content/mimiciv/3.0/ |

### Specification and cost used for running models

We made use of the top performing LLM to date, OpenAI's GPT-4o. Selection of model hyperparameters was in order to optimize performance. Temperature was set between 0.5 and 0.7.The maximum token limit was 100 tokens. According to OpenAI, a token is roughly equal to 4 characters and 100 tokens roughly equate to 75 words[14]. We found this was a generous allotment for GPT responses with 13 tokens per response on average. The token limit still served to prevent potentially excessive responses. On average, 241 input tokens were used but approached 600 tokens in some cases. In order to prime the GPT model to respond to these particular requests the following system prompt was used, "You are assisting in the standardization of clinical data dictionaries. Respond directly and concisely to each prompt, adhering to specified formats and contexts. Do not include labels, delimiters, or re-use the prompt in any way. Only the requested data". The system prompt was followed by 1 of 9 instructional prompts corresponding to different fields.An approximate cost breakdown of using GPT-4o to follow our methods detailed above is provided below.

| **Model** | **Input Token Cost** | **Output Token Cost** | **Avg. Input Tokens** | **Avg. Output Tokens** | **Approximate Cost Per Request + Response** |
|---|---|---|---|---|---|
| GPT-4o | $0.0050 / 1k tokens | $0.0150 / 1k tokens | 241 tokens | 13 tokens | $ 0.0014 |

### Example of Prompt Template Using Jinja2 Engine

Below is a prompt template used to collect potential aliases for a given concept to assist in mapping tasks. Jinja is a dynamic templating engine that uses control structure and expressions to insert the relevant data when available.

*Instructions: Based on the existing title for the data element below and the provided context, create a comma-separated list of appropriate alternative aliases, terms, and abbreviations commonly found in medical texts.*
*Biomedical term: ### {{ title }} ###*

*{% if additional_context %}*
*Additional context provided by the user:*
*{{ additional_context }}*
*{% endif %}*

*{% if selected_columns_data %}*

*Additional column data:*
*{% for col, value in selected_columns_data.items() %}*
*- {{ col }}: {{ value }}*
*{% endfor %}*
*{% endif %}*

### Detailed Interoperability Test Results for ADNI-ADSXLIST

The table below displays interoperability results for the ADNI ADSXLIST(Alzheimer's Disease Symptoms Checklist) data tables. The Header column contains the column headers from the table. The Title of Closest Match - GenCDE contains the title associated with a generated CDE that scored above the implicit threshold of the auto fuzziness parameter.

Completeness indicates the percentage of fields containing a non-null value for each column where a correct match was found. Compliance reports the percentage of non-null fields that contain expected value ranges, data types, and/or categorical values. Note that most columns in this table did not comply with our set standards. ADNI uses a coding convention where 1=Absent and 2=Present. Our permissible value set allows for 0=False/Absent and 1=True/Present as well as a "True" or "False" string. Using 1 and 2 to represent these values is not typical or recommended in forward-looking data science[15]. The columns that scored 100% compliance were categorized as 'Free Entry'.

The table is color coded to indicate which columns were mapped successfully. Red is an incorrect match while green is a correct match. This test was performed during the earlier stages of CDE repository development and prior to ingestion of aliases or the generation of new CDEs from ADNI materials.

| Header | Title of Closest Match - GenCDE | Completeness (%) | Compliance (%) | Source of GenCDE | ADNI Data Dictionary Definition |
|---|---|---|---|---|---|
| Phase | Cortical evoked potential steady-state time frequency analysis phase interval | x | x | NLM CDE Repository | ADNI Cohort ID |
| ID | Participant ID | x | x | CARD CDEs | Record ID |

| | | | | | |
|---|---|---|---|---|---|
| PTID | Participant ID | 100 | 100 | CARD CDEs | Participant ID |
| RID | Healthy Volunteer of PID | x | x | SNOMED CT | Participant roster ID |
| SITEID | Exostosis of Unspecified Site (ICD9, 726.91) | x | x | ICD | Site ID |
| VISCODE | Imaging aneurysm viscous dissipation measurement | x | x | NLM CDE Repository | Visit code |
| VISCODE2 | Scored Below Threshold for Any Potential Match | x | x | x | Translated visit code |
| VISDATE | Visit date | 100 | 0 | UPDRS 20n0153 | Registry EXAMDATE on matching VISCODE |
| USERDATE | Date of heaviest alcohol use | x | x | Populations Underrepresented in Mental illness Association Studies (PUMAS) | Date Record Created |
| USERDATE2 | Date of heaviest alcohol use | x | x | Populations Underrepresented in Mental illness Association Studies (PUMAS) | Date Record Last Updated |

| | | | | | |
|---|---|---|---|---|---|
| EXAMDATE | Cutaneous Candidiasis (ICD10, B37.2) | x | x | ICD | Examination Date |
| AXNAUSEA | Interview History of Nausea | 100 | 0 | CTDB Form Legend 00CH0134 | Nausea |
| AXVOMIT | Adverse Drug Reaction Question 16: Black Vomit | x | x | CTDB Form Legend 00CH0134 | Vomiting |
| AXDIARRH | Diarrhea | 100 | 0 | CTDB Form Legend 00CH0134 | Diarrhea |
| AXCONSTP | Cholecystitis | x | x | BTRIS Clinical Observations | Constipation |
| AXABDOMN | Abdominal Actinomycotic Infection (ICD10, A42.1) | x | x | ICD | Abdominal Pain |
| AXSWEATN | Anhidrosis (ICD10, L74.4) | 100 | 0 | ICD | Sweating |
| AXDIZZY | Becoming dizzy or falling | 100 | 0 | Populations Underrepresented in Mental illness Association Studies (PUMAS) | Dizziness |

| | | | | | |
|---|---|---|---|---|---|
| AXENERGY | Fatigue (feel tired or heavy, ache) | 100 | 0 | Neuropsychiatric | Low Energy |
| AXDROWSY | Glaucoma drops name PhenX, What is the name of the glaucoma drops you are using [PhenX], What is the name of the glaucoma drops you are using? | x | x | NLM CDE Repository | Drowsiness |
| AXVISION | Blurred Vision | 100 | 0 | BTRIS Clinical Observations | Blurred Vision |
| AXHDACHE | Headache | 100 | 0 | BTRIS Clinical Observations | Headache |
| AXDRYMTH | Systemic Lupus Eryth | x | x | BTRIS Clinical Observations | Dry Mouth |
| AXBREATH | Halitosis (ICD10, R19.6) | x | x | ICD | Shortness of Breath |
| AXCOUGH | Cough (ICD10, R05) | 100 | 0 | ICD | Coughing |
| AXPALPIT | Pale Optic Nerves | x | x | BTRIS Clinical Observations | Palpitations |

| | | | | | |
|---|---|---|---|---|---|
| AXCHEST | Acq Chest Deformity (ICD9, 738.3) | x | x | ICD | Chest Pains |
| AXURNDIS | Unilat Rad Neck Dissect (ICD9, 40.41) | x | x | ICD | Urinary discomfort (e.g., burning) |
| AXURNFRQ | Scored Below Threshold for Any Potential Match | x | x | x | Urinary frequency |
| AXANKLE | Ankle Swelling | 100 | 0 | BTRIS Clinical Observations | Ankle Swelling |
| AXMUSCLE | Other Plastic Operations on Muscle | x | x | BTRIS Clinical Observations | Musculoskeletal pain |
| AXRASH | Adverse Drug Reaction Question 1B: Presence of Skin Rash | x | x | CTDB Form Legend 00CH0134 | Rash |
| AXINSOMN | Insomnia, Unspecified (ICD10, G47.00) | 100 | 0 | ICD | Insomnia |
| AXDPMOOD | Stroke Specific Quality of Life Scale (SS-QOL) - interest mood scale | x | x | NLM CDE Repository | Depressed mood |

| | | | | | |
|---|---|---|---|---|---|
| AXCRYING | Excessive Crying Of Child, Adolescent, Or Adult (ICD10, R45.83) | 100 | 0 | ICD | Crying |
| AXELMOOD | Epiretinal membrane in the right eye. | x | x | BTRIS Clinical Observation | Elevated Mood |
| AXWANDER | Water Intake Measurement | x | x | CTDB Form Legend 00CH0134 | Wandering |
| AXFALL | Other and unspecified accidental fall | 100 | 0 | BTRIS Clinical Observation | Fall |
| AXOTHER | OTHER | OTHER | 100 | BTRIS Clinical Observations | Other/Diagnosis |
| AXSPECIF | COVID-19 Specific Medication Specify Other Type | x | x | NLM CDE Repository | If Other symptoms/diagnosis, specify: |
| update_stamp | Gastrostomy, Open; without Construction of Gastric Tube (eg, Stamm procedure) (CPT-HCPCS, 43830) | x | x | CPT-HCPCS | Update stamp |
